# Sequential mosaic variants in *KRAS* and *STAT5B* associated with a mixed phenotype of two acquired errors of immunity

**DOI:** 10.1101/2024.08.07.24311150

**Authors:** Julia Forkgen, Etienne Masle-Farquhar, Yves Fontaine, Amanda Russell, Shannon Ji, Timothy J Peters, Zhen Qiao, Michael Geaghan, Katherine JL Jackson, Jillian M Hammond, Ira W Deveson, Clementine David, Daniel A Lemberg, Nitin Gupta, Noemi Fuentes-Bolanos, Satu Mustjoki, Vivian Hwa, Stuart G Tangye, Paul E Gray, Owen M Siggs

## Abstract

Mosaic genetic variation has been implicated in the pathogenesis of both malignant and non-malignant immunological disease. Here, we report a unique case of postnatal acquisition of a gain-of-function (GoF) *KRAS* variant, with an additional GoF *STAT5B* variant, in a woman with inflammatory bowel disease, splenomegaly, thrombocytopenia, bronchiectasis, monocytosis, and eosinophilia. Targeted amplicon sequencing revealed widespread distribution of both variants in key immune cell populations, and in historical blood and tissue samples, with the emergence of both variants coinciding with the time of clinical presentation. Short- and long-read single cell RNA sequencing of patient cells highlighted a unique population of monocytes, with a broad distribution of both variants, and dysregulated cytokine signaling pathways. Flow cytometry revealed dysregulated STAT signaling, and the presence of a distinct population of highly granular CD24+ cells. Taken together with the clinical presentation, these findings led to a diagnosis of combined RAS-associated autoimmune leukoproliferative disorder (RALD) and non-clonal *STAT5B* GoF disease. To our knowledge, this is the first reported combination of two distinct acquired errors of immunity causing a mixed clinical phenotype, and highlights the importance of considering acquired monogenic diseases within a broader genomic context.

## Introduction

Mosaic variation is an increasingly recognised driver of non-malignant disease, particularly in disorders of inflammation and immunity^1^. Acquired gain-of-function (GoF) variants in the proto-oncogene *KRAS* have been reported in RAS-associated autoimmune leukoproliferative disease (RALD)^2,3^. RALD is not considered a malignancy, but is associated with increased risk of juvenile myelomonocytic leukemia (JMML)^4^.

Similarly, mosaic GoF variants in the Src homology 2 (SH2) domain of *STAT5B* have been identified not only in a non-malignant inflammatory disease, characterized by eosinophilia, atopic dermatitis, bronchiectasis and urticarial rash^5–7^, but also in individuals with immunodeficiency^8^, or with malignancies including T-cell large cell granular lymphocytic leukemia (T-LGLL)^9–11^, hepatosplenic T cell lymphoma^12^, and enteropathy associated intestinal T cell lymphoma (EATL)^13,14^.

Here, we report and characterize a novel case of co-occurring mosaic variants previously established to be GoF in *STAT5B* (T628S) and *KRAS* (G12S), in an individual presenting with clinical features of both monogenic acquired disorders.

## Methods

Detailed methods are described in the Supplemental Information. Briefly, whole genome sequencing was performed on blood- or tissue-derived DNA, and somatic variants identified by MuTect2^15^. Peripheral blood mononuclear cell (PBMC) populations were analyzed by flow cytometry, and/or purified by fluorescence-activated cell sorting (FACS). DNA extracted from sorted PBMCs, whole blood or archival biopsies underwent *STAT5B/KRAS* PCR amplicon sequencing. Single-cell mRNA sequencing (10X Chromium) was performed on samples multiplexed using TotalSeq (BioLegend) antibodies. A portion of 10X-generated pre-fragmented cDNA underwent poly-dT capture and long-read sequencing (Oxford Nanopore Technologies). STAT3/5 activation was measured by phosphoflow, following cytokine stimulation.

## Results and discussion

### Case presentation

A female patient initially presented with weight loss, diarrhea and abdominal pain, and was diagnosed with inflammatory bowel disease (IBD) resembling Crohn’s disease (Fig 1A). They later developed eosinophilic oesophagitis, recurrent infections, bronchiectasis with chronic right middle lobe collapse (Fig 1B), moderate to severe asthma and episodes of discharging otitis media. Splenomegaly and panhypogammaglobulinemia were both observed at different timepoints, with low IgG, absent IgA and IgM, and exceedingly low B cells and natural killer (NK) cells in the absence of immunosuppression (Fig 1C). Conversely, they presented with consistently elevated peripheral blood eosinophils and monocytes (Fig 1C). Gastrointestinal biopsies revealed infiltration of intraepithelial lymphocytes and eosinophils. The patient was treated with intravenous immunoglobulin (IVIG) and azathioprine at initial presentation, although azathioprine was later discontinued following concerns about possible treatment-related hematopoietic malignancy, in the context of a new anemia and thrombocytopenia.

**Figure 1.**
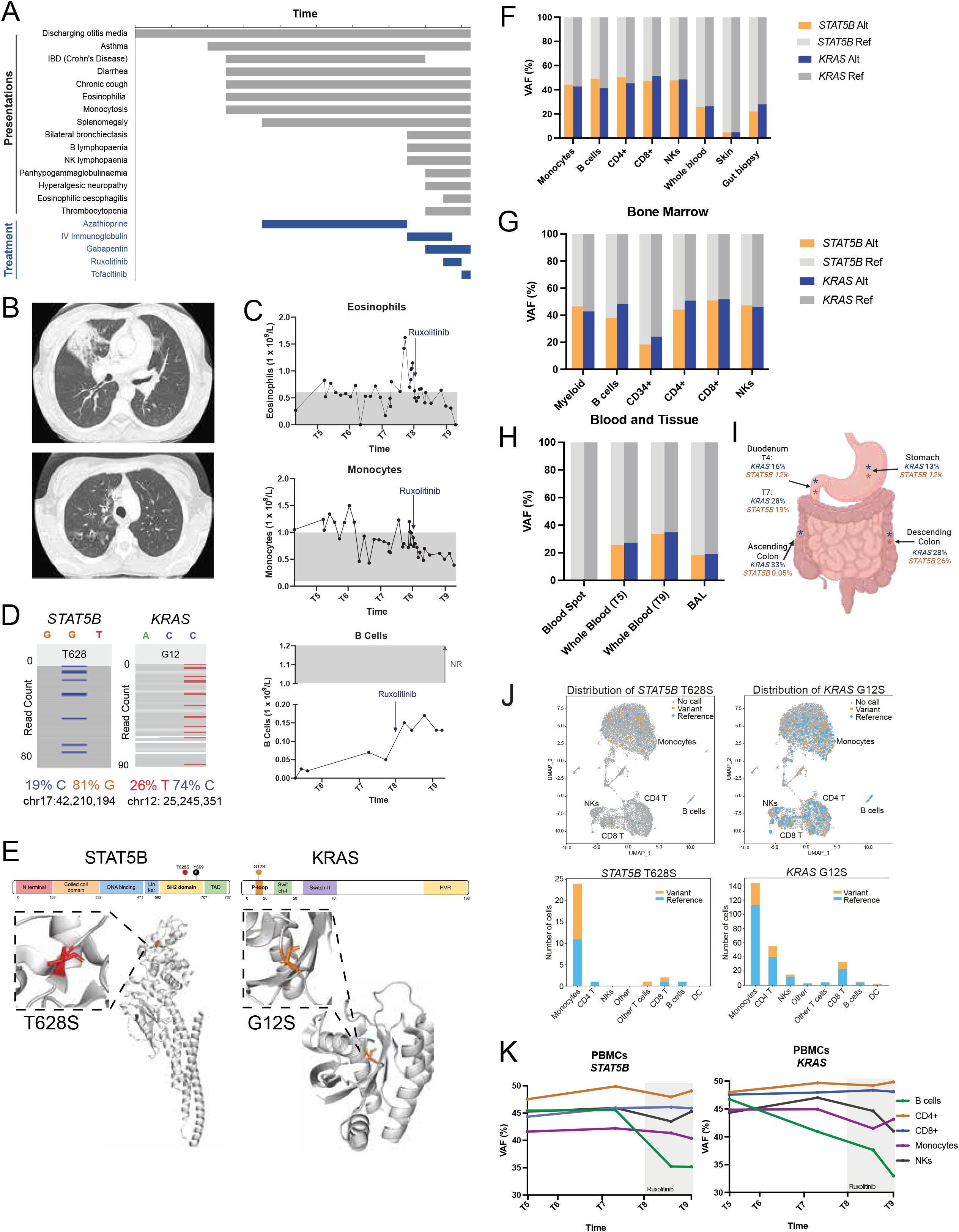
Clinical presentation and genetic investigations. **(A)** Chronology of clinical symptoms and treatment of the index patient. **(B)** Axial lung CT imaging displaying right middle lobe collapse (left image) and right upper lobe ground glass opacity and bronchiectatic change (right image). **(C)** Longitudinal eosinophil, monocyte and B cell counts. NR = Normal range **(D)** Mosaic variants identified in whole blood-derived DNA by whole genome sequencing. **(E)** Location and topology of missense mosaic variants. (**F-I)** *STAT5B* and *KRAS* VAFs in sorted leukocyte populations from blood **(F)**, bone marrow **(G)**, blood and tissue (**H)** and digestive tract tissue (**I). (J)** Seurat UMAP of nanopore long-read single-cell RNA sequencing data (top panel), overlaid with *STAT5B* (left) and *KRAS* (right) variant calls in single cells. Bottom panel, stacked bar plot of the total number of distinct reference and variant type cell barcodes for *STAT5B* and *KRAS* within each cell type annotation. (**K)** *STAT5B* and *KRAS* variant VAFs detected longitudinally in blood over a two year period (grey box indicates period of ruxolitinib treatment). T1 - T10 indicate various timepoints.

### Mosaic gain-of-function variants in STAT5B and KRAS

Diagnostic panel sequencing (Invitae Primary Immunodeficiency Panel) identified a single missense variant of uncertain significance in *STAT5B* (NM_012448.4:c.1883C>G, p.Thr628Ser, T628S, CADD 24.7). Although present at a very low frequency in gnomAD^16^ v4.1.0 (2 of 1,614,152 alleles), the variant had been reported in T-cell prolymphocytic leukemia^9^ and hepatosplenic T-cell lymphoma^12^, shown to be GoF and associated with increased proliferation and STAT5 phosphorylation^9^. Upon suspicion that this may represent an acquired variant, DNA was isolated from whole blood and a skin punch biopsy, and whole genome sequenced. DNA mutational signature analysis identified an azathioprine-associated signature (Supplemental Fig 1A-C). WGS confirmed the presence of *STAT5B* T628S in blood at a variant allele fraction (VAF) of 0.19 (Fig 1D, Supplemental Table 1). In contrast, this variant was not detected by capillary sequencing of skin and buccal swab from the patient, nor in blood from both parents. Genome-wide mosaic variant calling also revealed a blood-restricted variant in *KRAS* (NM_004985.5:c.34G>A, p.Gly12Ser, G12S), at a similar VAF of 0.26 (Fig 1D). While *KRAS* G12S was not in the original diagnostic panel, it is a well-established GoF cancer driver variant, including pancreatic cancer, multiple myeloma and intestinal T cell lymphoma^12,17^. Both variants were located in mutational hotspots within each gene (Fig 1E).

### Chronological, ontological and anatomical variant distribution

Amplicon-based sequencing of sorted leukocyte populations revealed *STAT5B* T628S and *KRAS* G12S variants in peripheral monocytes, B cells, natural killer (NK) cells, CD4 and CD8 T cells, at VAFs of 0.32-0.51 (Fig 1F-I). Both variants were detected in all tested sorted bone marrow populations, including CD34+ stem cells (*STAT5B* 0.18, *KRAS* 0.24 VAF)(Fig 1G). There was a 32% and 27% increase in *STAT5B* and *KRAS* VAFs in whole blood respectively, over a 2-year period (Fig 1H). Both variants were below the limit of detection in a blood spot taken at birth (Fig 1H), despite a sequencing coverage of 1.8M and 1.3M reads per *STAT5B* and *KRAS* variant respectively. In contrast, both variants were detected in multiple gastrointestinal and other tissue biopsies: in the descending colon at diagnosis prior to the commencement of azathioprine, in duodenal biopsies, the stomach and a bronchoalveolar lavage sample (Fig 1H-I). Only the *KRAS* variant however, was detected in an ascending colon biopsy (VAF = 0.33) This may suggest that clones harboring the *KRAS* variant alone, or both variants, populated different inflammatory infiltrates at different times. Alternate possibilities include clones harboring a single variant each, or a somatic reversion of the *STAT5B* variant. However, under the assumption that both variants are heterozygous, the observation that most samples have a VAF of >0.25 would mean that >50% of cells are heterozygous for both variants, and therefore most likely to have occurred sequentially.

Nanopore long-read single-cell RNA sequencing identified all major immune lineages (Fig 1J) and allowed transcript-based genotyping of individual cells. Consistent with sorted blood cells, this revealed widespread distribution of both *STAT5B* and *KRAS* variants across all leukocyte populations, with 16/30 [53%] and 63/262 [24%] cells genotyped for each respective variant (Fig 1J). Due to limited sequencing depth, no cells were genotyped at both variant positions.

Treatment with the JAK inhibitor ruxolitinib (5 mg b.d.) occurred following identification of the *STAT5B* variant. This resulted in reduced eosinophils and monocytes, and increased B cells (Fig 1C). There was little change in the VAF of either variant in most leukocyte populations following treatment, with the exception of reduced VAFs in B cells (Fig 1K), which in the context of increased B cell numbers would be consistent with an expansion of B cells without either GoF variant^9,18,19^.

### Absence of malignancy, with functional evidence of dysregulated signal transduction

Given the association of both variants with hematological malignancy, circulating lymphocytes were investigated by flow cytometry and single cell RNA sequencing. Most apparent was a ∼5-fold increase in percentages of high granularity (SSC^high^), CD14+ monocytes, and ∼30-fold higher SSC^high^ CD20- CD14- CD56- CD16- CD3- CD24+ cells, at every time point (Fig 2A). Due to the lack of expression of CD14, CD4 or CD16 this population did not correspond to classical monocytes, monoblasts or atypical monocytes, although CD24 expression in this context has previously been described as a highly specific marker of monocytic leukemia^20^. Single-cell RNA sequencing also revealed an expanded monocyte-annotated population in the patient PBMCs, which clustered separately to monocyte populations from three healthy donors and four disease controls (Fig 2C). Despite this, JMML was not diagnosed on bone marrow aspirate, although due to the significant risk of hematological malignancy, they were kept under continued surveillance, with ongoing consideration for early allogeneic hematopoietic stem cell transplantation.

**Figure 2.**
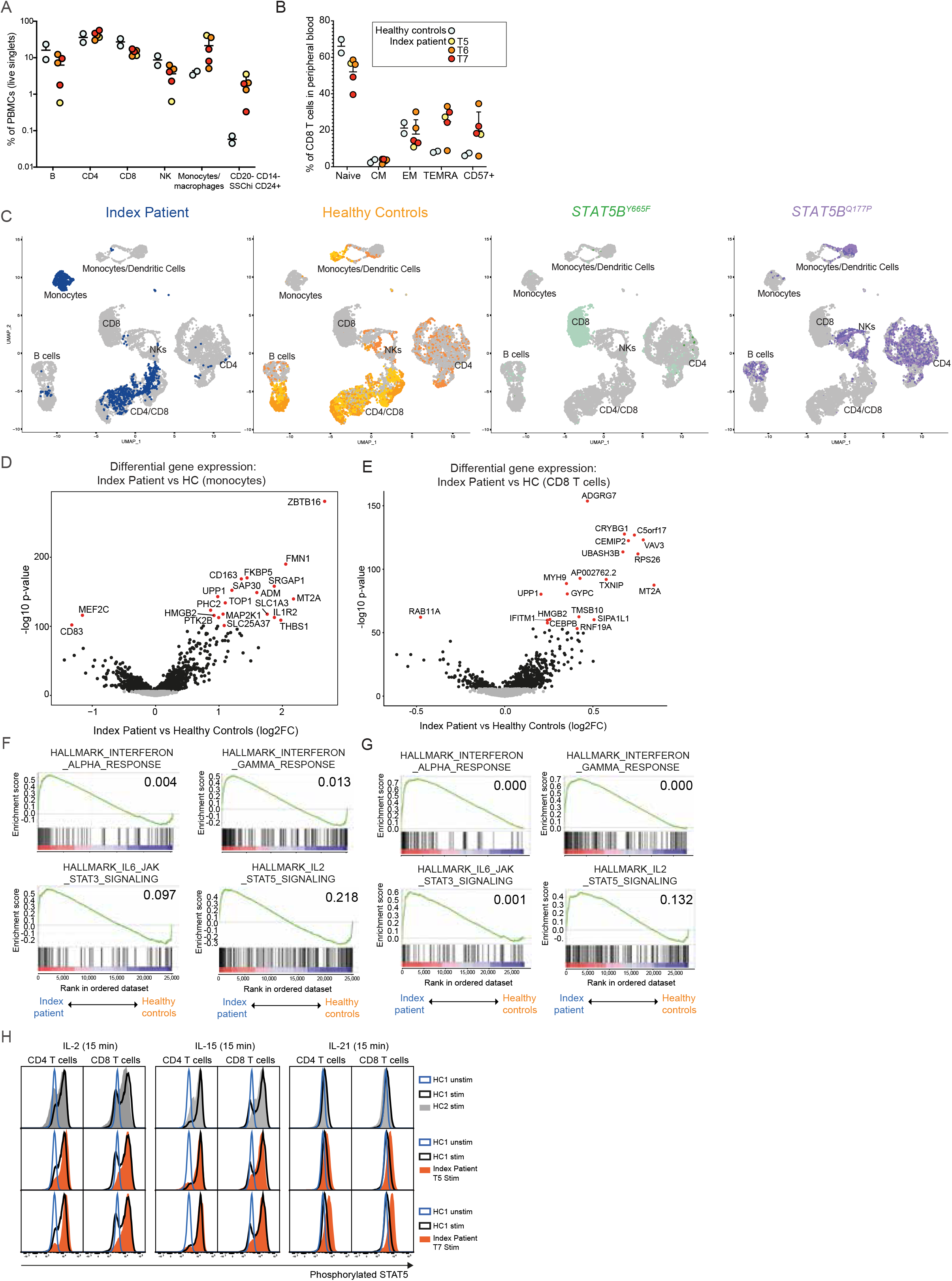
Functional and transcriptional impact and cell-type distribution of mosaic variants. **(A)** Percentage of B, CD4 or CD8 T, NK, monocyte/macrophage, or CD20- CD14- SSC++ CD24++ cells, within live singlet PBMCs, as measured by flow cytometric analysis. **(B)** Percentage of CD45RA+ CCR7+ naive, CD45RA- CCR7+ central memory (CM), CD45RA- CCR7- effector memory (EM) and CD45RA+ CCR7- terminally differentiated effector memory expressing CD45RA (T-EMRA) CD8 T cell populations within PBMCs of the index patient at three timepoints, relative to age-matched healthy controls. **(C)** UMAPs following scRNAseq of PBMCs and dimensionality reduction analysis, highlighting index (left; blue), healthy control (middle-left; orange), T-LGLL (middle-right; green) or STAT5B LoF (right; purple) cells, and all other projected cells in gray. **(D**,**E)** Volcano plots of log2 expression fold-change (log2FC) versus moderated t-statistic for differentially expressed genes in patient relative to healthy control monocytes **(D)** or CD8 T cells **(E)**. Red symbols indicate genes below an arbitrary, highly significant corrected p-value threshold. **(F**,**G)** Gene set enrichment analysis (GSEA) enrichment scores (y axis) for rank-ordered genes (x axis) differentially expressed by index relative to healthy control cells, for the most statistically significant (and cytokine/IFN-related) hallmark gene ontologies, for monocytes (**F)** and CD8 T cells (**G)**. Values represent family-wise error rate adjusted (FWER) p-values (<0.05). **(H)** Phosphoflow analysis showing STAT5 phosphorylation in response to stimulation (stim) with IL-2 (left), IL-15 (middle), or IL-21 (right), in index patient at two timepoints, relative to representative healthy control. UMAPs= uniform manifold approximation and projection, scRNAseq = single cell RNA sequencing, T-LGLL = T cell large granular lymphocytic leukemia, T1-T10 indicate various timepoints.

Flow cytometric analysis also revealed an increase in peripheral CD57+ CD8 T cells (Fig 2B). Despite this, the patient displayed no evidence of T-LGLL on the basis of a polyclonal TCR Vβ repertoire, and scRNAseq-based clustering showing that the patient’s CD8 T cells cluster distinctly to those of 2 patients with CD8 T-LGLL, both harboring the *STAT5B* GoF variant Y665F, and cells harboring a dominant negative *STAT5B* variant (Q177P) (Fig 2C, Supplemental Fig 2).

Relative to healthy controls, the patient’s monocytes or CD8 T cells had significantly increased expression of 771 and 698 genes, and decreased expression of 714 and 201 genes, respectively (Fig 2D-E, Supplemental Table 2-9). Consistent with the GoF nature of both variants, pathway analyses in monocytes and CD8 T cells revealed statistically significant dysregulation of transcripts associated with responses to IFNα and IFNγ, IL-6/JAK/STAT3 signaling, and IL2/STAT5B signaling (FDR q <0.05), though not all pathways were significant when family-wise error rate adjustment was applied (PWER p values) (Fig 2F-G, Supplemental Fig 3, Supplemental Table 10a-d, 11a-d).

Further, relative to two healthy controls, T cells collected from the patient at two timepoints had increased STAT5B phosphorylation in response to stimulation with IL-21 (Fig 2H). IL-21 primarily stimulates STAT3, although it can also activate STAT5, and in the context of this patient may reflect an exaggeration of STAT5B activation secondary to *STAT5B* T628S and/or *KRAS* G12S.

### Combinatorial phenotypes of acquired variants

*STAT5B* GoF variants including T628S and the sister variant N642H have previously been associated with non-clonal eosinophilia^5–7^, lymphocyte-variant hypereosinophilia^21^ and recurrent infiltration of eosinophils in the gastrointestinal tract ^5^. Chronic monocytosis, splenomegaly, cytopenia and recurrent infections including bronchiectasis and otitis media are strongly associated with RALD^2–4^, all of which were observed in this patient. Based on our findings, the patient was re-classified as a combined clinical entity of RALD and non-clonal *STAT5B* GoF disease ^3,5,6^ (Supplemental Table 12), and commenced on ruxolitinib with subsequent improvements in splenomegaly, bronchiectasis, and quality of life.

While acquired variants have recently been shown to act combinatorially to cause disease^22^, to our knowledge this is one of the first reported combinations of two distinct acquired errors of immunity. Similar to malignancy, these cases highlight the importance of considering non-malignant acquired genetic diseases within a broader genomic context.

## Data Availability

All data produced in the present study are available upon reasonable request to the authors

## Acknowledgements

This study was supported by a Snow Medical Fellowship (to OMS). SGT is supported by an Investigator Grant awarded by the National Health and Medical Research Council (NHMRC) of Australia (Level 3; 1176665).

## References

1. Mustjoki S, Young NS. Somatic Mutations in “Benign” Disease. N. Engl. J. Med. 2021;384(21):2039–2052.

2. Niemela JE, Lu L, Fleisher TA, et al. Somatic KRAS mutations associated with a human nonmalignant syndrome of autoimmunity and abnormal leukocyte homeostasis. Blood. 2011;117(10):2883–2886.

3. Neven Q, Boulanger C, Bruwier A, et al. Clinical Spectrum of Ras-Associated Autoimmune Leukoproliferative Disorder (RALD). J. Clin. Immunol. 2021;41(1):51–58.

4. Calvo KR, Price S, Braylan RC, et al. JMML and RALD (Ras-associated autoimmune leukoproliferative disorder): common genetic etiology yet clinically distinct entities. Blood. 2015;125(18):2753–2758.

5. Ma CA, Xi L, Cauff B, et al. Somatic STAT5b gain-of-function mutations in early onset nonclonal eosinophilia, urticaria, dermatitis, and diarrhea. Blood. 2017;129(5):650–653.

6. Eisenberg R, Gans MD, Leahy TR, et al. JAK inhibition in early-onset somatic, nonclonal STAT5B gain-of-function disease. J. Allergy Clin. Immunol. Pract. 2021;9(2):1008–1010.e2.

7. Ding F, Wu C, Li Y, et al. A case of hypereosinophilic syndrome with STAT5b N642H mutation. Oxf Med Case Reports. 2021;2021(1):omaa129.

8. Savola P, Martelius T, Kankainen M, et al. Somatic mutations and T-cell clonality in patients with immunodeficiency. Haematologica. 2020;105(12):2757–2768.

9. Kiel MJ, Velusamy T, Rolland D, et al. Integrated genomic sequencing reveals mutational landscape of T-cell prolymphocytic leukemia. Blood. 2014;124(9):1460–1472.

10. Andersson EI, Tanahashi T, Sekiguchi N, et al. High incidence of activating STAT5B mutations in CD4-positive T-cell large granular lymphocyte leukemia. Blood. 2016;128(20):2465–2468.

11. Bhattacharya D, Teramo A, Gasparini VR, et al. Identification of novel STAT5B mutations and characterization of TCRβ signatures in CD4+ T-cell large granular lymphocyte leukemia. Blood Cancer J. 2022;12(2):31.

12. McKinney M, Moffitt AB, Gaulard P, et al. The Genetic Basis of Hepatosplenic T-cell Lymphoma. Cancer Discov. 2017;7(4):369–379.

13. Moffitt AB, Ondrejka SL, McKinney M, et al. Enteropathy-associated T cell lymphoma subtypes are characterized by loss of function of SETD2. J. Exp. Med. 2017;214(5):1371–1386.

14. Roberti A, Dobay MP, Bisig B, et al. Type II enteropathy-associated T-cell lymphoma features a unique genomic profile with highly recurrent SETD2 alterations. Nat. Commun. 2016;7:12602.

15. Benjamin D, Sato T, Cibulskis K, et al. Calling Somatic SNVs and Indels with Mutect2. bioRxiv. 2019;861054.

16. Chen S, Francioli LC, Goodrich JK, et al. A genomic mutational constraint map using variation in 76,156 human genomes. Nature. 2023;

17. Cook JH, Melloni GEM, Gulhan DC, Park PJ, Haigis KM. The origins and genetic interactions of KRAS mutations are allele-and tissue-specific. Nat. Commun. 2021;12(1):1808.

18. Hyrenius-Wittsten A, Pilheden M, Sturesson H, et al. De novo activating mutations drive clonal evolution and enhance clonal fitness in KMT2A-rearranged leukemia. Nat. Commun. 2018;9(1):1770.

19. Oshima K, Khiabanian H, da Silva-Almeida AC, et al. Mutational landscape, clonal evolution patterns, and role of RAS mutations in relapsed acute lymphoblastic leukemia. Proc. Natl. Acad. Sci. U. S. A. 2016;113(40):11306–11311.

20. Raife TJ, Lager DJ, Kemp JD, Dick FR. Expression of CD24 (BA-1) predicts monocytic lineage in acute myeloid leukemia. Am. J. Clin. Pathol. 1994;101(3):296–299.

21. Umrau K, Naganuma K, Gao Q, et al. Activating STAT5B mutations can cause both primary hypereosinophilia and lymphocyte-variant hypereosinophilia. Leuk. Lymphoma. 2023;64(1):238–241.

22. De Langhe E, Van Loo S, Malengier-Devlies B, et al. TET2-Driver and NLRC4-Passenger Variants in Adult-Onset Autoinflammation. N. Engl. J. Med. 2023;388(17):1626–1629.

