## Supplemental Data for "Sequential mosaic variants in *KRAS* and *STAT5B* associated with a mixed phenotype of two acquired errors of immunity"

#### Supplemental Materials and Methods

##### *Participants and sample processing*

Whole blood was collected by venipuncture in K-EDTA or ACD collection tubes, and plasma isolated by centrifugation of ACD vials at 800 g for 10 min at room temperature. Peripheral blood mononuclear cells (PBMCs), or mononuclear cells from a bone marrow aspirate, were isolated by Ficoll paque density gradient centrifugation. Fresh and archival gut biopsies, bronchoalveolar lavage and a skin punch biopsy were also collected from the index patient. Additional samples were obtained from patients with autosomal dominant growth hormone insensitivity (germline *STAT5B* Q177P variant) and CD8 T-LGLL (mosaic *STAT5B* Y665F). This study was approved by the Sydney Local Health District Ethics Review Committee (protocols X20-0177 and 2020/ETH00998, X16-0210 and 2019/ETH06359), Western Sydney Local Health District Human Research Ethics Committee (HREC/17/WMEAD/551), Cincinnati Children's Hospital Medical Center (GBGD-2014-5919) and Helsinki University Central Hospital Ethics Committee (Helsinki, Finland)[23]. Appropriate written informed consent was obtained from all participants or their guardians.

##### *Whole genome sequencing and somatic variant calling*

DNA was extracted from whole blood and skin punch biopsy samples of the index patient using the DNeasy Blood and Tissue Kit (Qiagen). Whole genome sequencing (30X Illumina PCR-Free) was performed on an Illumina NovaSeq 6000 instrument, with FASTQ outputs aligned to the GRCh38 reference. MuTect2[15] was run in tumour-normal mode to identify somatic variants present in blood-derived DNA, but absent from skin-derived DNA.

##### *Mutational signature analysis*

All putative blood-restricted somatic variants identified by the MuTect2 analysis (PASS only) were subjected to mutational signature deconvolution using the MutationalPatterns R package[24]. Variants were limited to single nucleotide variants for the analysis. Variants were fitted to mutational signatures from COSMIC (v3.2) strictly with a maximum delta value of 0.002. The signature contributions were plotted using the absolute number of variants. A mutational spectrum showing the relative contributions of base substitutions types was plotted, with a distinction made between C>T substitutions at CpG sites and other sites. A 96 trinucleotide profile was also made showing the relative contributions of each base substitution type including the sequence context one base 5' and one base 3' of the mutated base.

##### *Flow cytometry*

Cryopreserved PBMCs were thawed, washed and resuspended in cold sterile PBS containing 2% heat inactivated Fetal Calf Serum (FCS). Cells were transferred into 96-well round-bottom plates, and 1-3 x $10^6$ cells per well were incubated for 30 min at 4C in PBS 2% FCS containing fluorophore-conjugated antibodies (see Supplemental Table 13). Following washes, cells were washed and resuspended in PBS for acquisition on a BD Fortessa or BD Aria III. To bulk-sort leukocyte populations, antibody-stained cells were left unfixed and resuspended in PBS 2% FCS for acquisition and FACS using the BD Aria III. To measure levels of phosphorylated STAT5 and STAT3, PBMCs were thawed as above and plated with 5-7.5 x $10^5$ cells per well, followed by stimulation with IL2, IL15 and IL21 cytokines (100 ng/ml) for 15 min, followed by fixation in 50 uL of ice-cold 100% methanol for 30 min at 4C. Cells were washed three times by addition of ice-cold PBS 2% FCS, centrifugation and aspiration of supernatant. Following washes, cells were incubated for 30 min at 4C with PBS 2% FCS containing fluorophore-conjugated antibodies (see Supplemental Table 13). Cells were then washed and resuspended in 4C PBS and acquired on an LSRII SORP instrument.

***Amplicon-based sequencing***
PBMCs and mononuclear bone marrow cells were bulk-sorted by Fluorescence-Activated Cell Sorting (FACS) into Eppendorf tubes containing cold sterile PBS 5% FCS, using canonical markers for CD4 and CD8 T cells, B cells, monocytes, natural killer cells and in the case of bone marrow, CD34+. DNA was subsequently extracted from monocyte, natural killer cells and CD34+ cells using the QIAamp DNA Blood Kit (Micro) (Qiagen), whilst DNA and RNA were extracted from B cell, CD4 and CD8 T cell populations using the AllPrep DNA/RNA Micro kit (Qiagen). The QIAamp DNA Blood Kit (Mini) was used to extract DNA from a Guthrie card blood spot and BAL scraping. To extract DNA from paraffin-embedded archival gut biopsies, tissue was macro-dissected from the paraffin block, and incubated with deparaffinization solution (Qiagen) for 3 min at 56ºC, followed by the addition of Buffer ATL and an overnight incubation with Proteinase K at 56ºC. DNeasy blood and Tissue kit (Qiagen) was subsequently used to extract DNA. To identify variants of interest, oligonucleotide primers (IDT) were designed to capture *STAT5B* variants T628S, Q177P and Y665F and *KRAS* G12S **(**Supplemental Table 14**)**, and barcoded to allow read identification following pooling. PCR was performed to amplify variants in DNA from sorted leukocyte populations, using Q5 2x High Fidelity Mastermix (New England Biolabs), primers (0.5uM) as per Table X, and 2ul input DNA (~2-4ng), with 35 cycles of amplification performed: 98ºC 10s, 60ºC (*STAT5B* Y665F, *KRAS* G12S) or 64ºC (*STAT5B* T628S, *STAT5B* Q177P) 30s, 72C 40s. Following purification of PCR products using AMPure XP Beads (Agencourt) as per manufacturer guidelines, complete adaptor sequences and index barcodes were added using the primers from the Illumina Nextera Index Kit (Illumina). Five cycles of PCR amplification were subsequently performed using Q5 2X High Fidelity Mastermix (New England Biolabs); 98ºC 10s, 63ºC 30s, 70ºC 3 min. Product was purified using AMPure XP Beads (Agencourt), and dsDNA concentration quantified using a Quant-iT Picogreen assay (Invitrogen). To pool samples, the amplified *STAT5B* and *KRAS* PCR products from the index patient each accounted for 1/3rd of the final sample concentration to achieve greater sequencing depth, whilst the remaining third of the submitted sample consisted of PCR products from the index and control patients pooled at equimolar concentrations. Samples were sequenced using the MiSeq v2 2 x 250 bp sequencing kit (Illumina), performed by Ramaciotti Centre for Genomics (UNSW Sydney, Australia).

FASTQ files for paired end sequencing libraries were aligned to reference amplicon gene sequences retrieved from GRCh38 using the Burrows Wheeler Aligner (BWA, version 0.7.17-r118) sampe option to generate paired-end alignments[25] Alignments were converted to bam files, sorted and indexed using Samtools (version 1.17)[26] before generating a variant call files (VCF) using the mpileup and call functions of Bcftools (version 1.16)[26]. The variant allele frequencies per sample were subsequently calculated from parsing each VCF with the vcfR package [27] and summarising with the tidyverse package[28] in R.

***Single-cell RNA sequencing***
Cells were prepared for single-cell RNA sequencing as described previously[29]. Briefly, PBMCs were thawed as described earlier, resuspended in cold sterile PBS 10% FCS and incubated for 20 min at 4°C with TotalSeq™ DNA-barcoded anti-human ‘Hashing’ antibodies (BioLegend) at a 1/100 final dilution (Supplemental Table 15). The TotalSeq-A™ antibodies conjugated to DNA oligonucleotides stained all leukocytes in the relevant samples. During the incubation, cells were transferred into a 96-well round bottom plate, on ice. Following incubation, cells were washed three times in cold PBS 2% FCS and the hashed populations pooled into a mixture. Following washes, the cell mixture was resuspended in cold PBS 5% FCS, for single-cell RNA sequencing using the 10X Genomics platform. The Garvan-Weizmann Centre for Cellular Genomics (GWCCG) performed the 10X capture and sequencing of resulting cDNA samples, using the Chromium Next GEM Single Cell 3' Reagent Kit v3.1 Dual Index (10X Genomics) as per the manufacturer protocol. A total of 25 000 cells were captured per reaction, using the Illumina S4 flow cell.

RNA libraries were sequenced on an Illumina NovaSeq 6000 (NovaSeq Control Software v 1.6.0 / Real Time Analysis v3.4.4) using a NovaSeq S4 230 cycle kit (Illumina, 20447086) as follows: 28bp (Read 1), 90bp (Read 2) and 10bp (Index). Sequencing generated raw data files in binary base call (BCL) format. These files were demultiplexed and converted to FASTQ using Illumina Conversion Software (bcl2fastq v2.19.0.316). Alignment, filtering, barcode counting and UMI counting were performed using the Cell Ranger Single Cell Software v6.0.2 (10X Genomics). Reads were aligned to the GRCh38-2020-A human (GENCODE v32/ Ensembl 98) reference genome. Hashtag-oligo counts (HTO) were processed with CITE-seq-Count (version 1.4.5, github.com/Hoohm/CITE-seq-Count).

*Cell clustering and annotation*
Count matrices for gene expression (from Cell Ranger) and HTO and CITE-seq antibodies (from CITE-seq-Count) were imported into R version 4.2.2 (2022-10-31)[30] using the Seurat v4 (version 4.3.0) package[31]. Using Seurat, HTO counts were normalised by centered log ratio (CLR) transformation and cells were demultiplexed to samples using the HTODemux function. Sample identities for individual cells were exported to a comma-separated (csv) file. Gene expression count data was filtered for mitochondrial percentage (>20%), library size and feature count using scater (version 1.24.0)[32]. Only cells with barcodes for gene expression, demultiplexed HTO and CITE-seq were retained. A Seurat object was generated with two data slots; RNA (for gene expression count data) and ADT (for CITE-seq count data). HTO information for each cell barcode was imported from csv using tidyverse (version 2.0.0)[28], and added to the Seurat object using AddMetaData. RNA data was normalised and scaled with SCTransform (SCT) and ADT data was normalised by CLR. Principal component analysis was run on the SCT counts for 50 dimensions and an Elbow plot was used to select the number of dimensions (ndims = 30) for dimensionality reduction via Uniform Manifold Approximation and Projection (UMAP). To examine the impact of TCR and immunoglobulin VDJ genes on clustering, a version of the processing was also performed following the removal of TR and IG VDJ genes from gene expression count matrix. Cells were clustered by the shared nearest neighbour (SNN) based algorithm implemented by Seurat's FindClusters function. This was run at a range of resolutions. To assist with cluster labelling, Azimuth[33] was applied to the data using the human PBMC reference set to provide cell type predictions for individual cells and gene markers for each cluster were defined.

Gene markers, or differentially expressed genes (DGE), were explored in two ways. First, each cluster was compared to all other cells using Seurat's FindAllMarkers. Second, pairwise comparisons between all pairs of clusters was performed using FindMarkers. After clustering analysis, supervised differential expression between individuals was performed. Single cell gene expression counts were normalised using Sanity[34] with default values. Differentially expressed genes were calculated using limma[35] on log expression quotients using a multifactor linear model, where each comparison was formulated as a pairwise contrast. Bonferroni correction was applied to each set of p-values and statistically significant differentially expressed genes were defined as having a family-wise error rate (FWER) < 0.05. Gene set enrichment analysis (GSEA)[36] was performed with 10,000 permutations on the Hallmark (h) ontology.

***Long read single-cell RNA sequencing***
Cells from the index patient were prepared for single-cell RNA sequencing using a Next GEM Single Cell 3' Kit (V3.1, 10X Genomics), as described above. Following full-length cDNA 10X capture generation, 10ng of the cDNA library was used as input for long read sequencing. Libraries were prepared following the 3' workflow from ONT ("Single-cell transcriptomics with 3' cDNA prepared using 10X Genomics on PromethION") using the SQK-LSK114 kit (Oxford Nanopore). The prepared library (30ng) was loaded on a PromethION flow cell (FLO-PRO114M) and sequenced for 72 hours. The raw data was base-called using dorado v7.1.4 to generate a FASTQ file. The wf-single-cell pipeline (https://github.com/epi2me-labs/wf-single-cell) was applied to the FASTQ, using GRCh38 as

a reference genome, to generate a BAM file containing reads with their assigned barcodes and UMIs, their alignments to the reference genome, and a matrix of read counts per cell. PCA was performed on the gene expression matrix using the function RunPCA in Seurat, and the first 30 principle components were used to generate a UMAP in R using the Seurat package [31]. The UMAP clusters were annotated with cell lineage identities using the Azimuth package with the reference 'pbmcref', at the annotation resolution level 'L1'.

The pysam package in Python was used to generate variant calls on long-read scRNAseq data. For both *STAT5B* and *KRAS*, the fetch function was used to iterate over each read that (i) was labelled as belonging to the gene of interest and (ii) explicitly had a nucleotide aligned to the variant site. The identity of the nucleotide at that location determined whether the read was annotated as a wild type or mutant allele. Reads corresponding to neither the wild-type reference nor the variant nucleotide change were assumed to result from read errors, and were thus excluded. As there were at most two reads for each of our genes of interest assigned to each unique cell barcode, each cell barcode was labelled as either alternate or reference for each of the genes of interest. UMAP coordinates were subsequently exported from Seurat and the matplotlib package in Python[37] used to overlay mutational statuses of the cell barcodes onto the UMAP. Similarly, matplotlib was used to overlay wild type and mutant type cell barcodes per cell type, onto the Azimuth labelled UMAP plot.

**Supplemental Tables**

**Supplemental Table 1. List of somatic coding variants identified in index patient blood, using WGS.** Candidate disease-associated variants are bolded.

| SYMBOL | CHROM | POS | REF | ALT | HGVSp |
|---|---|---|---|---|---|
| *HS6ST1* | chr2 | 128268653 | G | T | ENSP00000259241.6:p.Arg249Ser |
| *HS6ST1* | chr2 | 128268675 | G | A | ENSP00000259241.6:p.Cys241= |
| *PDCD11* | chr10 | 103423619 | T | G | ENSP00000358812.3:p.Val908= |
| *UBC* | chr12 | 124911831 | A | G | ENSP00000344818.5:p.Asp647= |
| ***KRAS*** | **chr12** | **25245351** | **C** | **T** | **ENSP00000256078.5:p.Gly12Ser** |
| *CD163L1* | chr12 | 7432645 | T | C | ENSP00000315945.3:p.Ile179Met |
| *CD163L1* | chr12 | 7432647 | T | C | ENSP00000315945.3:p.Ile179Val |
| *AHNAK2* | chr14 | 104943913 | G | C | ENSP00000353114.4:p.Leu3846Leu |
| *AHNAK2* | chr14 | 104945428 | G | T | ENSP00000353114.4:p.Asp3341Glu |
| *IGHV4-4* | chr14 | 106012171 | C | T | ENSP00000410711.2:p.Gly35Glu |
| ***STAT5B*** | **chr17** | **42210194** | **G** | **C** | **ENSP00000293328.3:p.Thr628Ser** |
| *PLIN4* | chr19 | 4511329 | T | C | ENSP00000301286.3:p.Lys863= |

= Represents synonymous SNPs

**Supplemental Table 12. Clinical features observed in non-clonal *STAT5B* gain of function disease, RALD, and the current case.** Features observed in the current case are bolded.

| | **Non-clonal *STAT5B* GoF disease** | **RALD** | **Current case** |
|---|---|---|---|
| **Non-clonal eosinophilia** | 100% (4/4 cases)[5–7] | NA | Observed |
| **Recurrent infections, including bronchitis, otitis media** | 75% (¾ cases) [5,6] | 24% (7/29) [3,38,39] | Observed |
| Urticarial skin rash | 75% (¾ cases)[5,6] | NA | NA |
| Atopic dermatitis | 75% (¾ cases) [5,6] | NA | NA |
| Elevated IgE | 67% (2/3 cases) [5,6] | NA | NA |
| **Diarrhea** | 50% (2/4 cases)[5,6] | 3% (1/31)[39] | Observed |
| **Gastrointestinal complications** | 50% (2/4) [5,6] | 13% (4/31)[3]<br>(All 4 cases carried *KRAS* mutations) | Observed |
| **Anemia** | 25% (1/4) [6] | 13% (4/31)[3,38–40] | Observed |

| | | | |
|---|---|---|---|
| **Splenomegaly** | 25% (¼ cases)[6]<br>Eisenberg 1/1 | 97% (32/33)[3,38–43] | Observed |
| Chronic lymphadenopathy | 25% (1/4)[6] | 89% (17/19)[3,38,39] | NA |
| **Cytopenias, including thrombocytopenia** | 25% (1/4) [7] | 84% (16/19)[3,38,40,43] | Observed |
| Hypergammaglobulinemia | NA | 71% (20/28)[3,39,42] | NA |
| **Hypogammaglobulinemia** | NA | 3% (1/31)[39]<br>Hypergammaglobulinemia which shifted to hypogammaglobulinemia | Observed |
| **Asthma** | NA | NA | Observed |
| **Regional pain syndrome** | NA | NA | Observed |

**Supplemental Table 13. Antibodies used for flow cytometry and FACS.**

| **Antibody target** | **Fluorophore** | **Provider** | **Catalogue #** | **Clone** | **RRID** |
|---|---|---|---|---|---|
| pSTAT5 | AF488 | BD BioSciences | 612598 | Clone 47/Stat5(pY694) | AB_399881 |
| pSTAT3 | PerCP Cy5.5 | BD BioSciences | 560114 | Clone 4/P-STAT3 | AB_1645335 |
| CD16 | PE | BD BioSciences | 347617 | B73.1 | AB_400331 |
| CD56 | PE | BD BioSciences | 347747 | MY31 | AB_400346 |
| CD57 | APC | BD BioSciences | 560845 | NK-1 | AB_10563760 |
| CD19 | APC Cy7 | BD BioSciences | 557791 | SJ25C1 | AB_396873 |
| CD3 | BV421 | BD BioSciences | 562426 | UCHT1 | AB_11152082 |
| Zombie Aqua | N/A | BioLegend | 423101 | N/A | AB_11124344 |
| CD45RA | BV605 | BD BioSciences | 562886 | HI100 | AB_2737865 |
| CD20 | BV650 | BD BioSciences | 563780 | 2H7 | AB_2744327 |
| CD14 | BV650 | BD BioSciences | 563419 | M5E2 | AB_2744286 |
| CD27 | BV786 | BD BioSciences | 563327 | L128 | AB_2744353 |
| CD8 | BUV395 | BD BioSciences | 563795 | RPA-T8 | AB_2722501 |

| | | | | | |
|---|---|---|---|---|---|
| CD4 | BUV737 | BD BioSciences | 564305 | SK3 (also known as Leu3a) | AB_2713927 |
| CD19 | BB515 | BD BioSciences | 564457 | HIB19 | AB_2744309 |
| CD38 | PerCP Cy5.5 | BD BioSciences | 551400 | HIT2 | AB_394184 |
| CD23 | PE Cy7 | eBioscience | 25-0238-42 | EBVCS2 | AB_1907366 |
| CCR7 | PE Cy7 | BD BioSciences | 557648 | 3D12 | AB_396765 |
| CD3 | APC Cy7 | BD BioSciences | 557832 | SK7 (also known as Leu-4) | AB_396890 |
| CD24 | BV421 | BD BioSciences | 562789 | ML5 | AB_2737796 |
| CD11c | BV786 | BD BioSciences | 740966 | B-ly6 | AB_2740591 |

**Supplemental Table 14. Primers used for amplicon sequencing.**

| Variant | Forward Primer Barcode (5'-3') | Forward Primer (5'-3') | Reverse Primer Barcode (3'-5') | Reverse Primer (3'-5') |
|---|---|---|---|---|
| *KRAS* G12S | CGTCGGCAGCGTCAGATGTGTATAAGAGACAG | ATGCATATTAAAACAAGATTTACCTC | GTCTCGTGGGCTCGGAGATGTGTATAAGAGACAG | AGTGTATTAACCTTATGTGTGACATG |
| *STAT5B* T628S | TCGTCGGCAGCGTCAGATGTGTATAAGAGACAG | GTTTGTAAACAAGCAACAGGC | GTCTCGTGGGCTCGGAGATGTGTATAAGAGACAG | CTGACCTCAACAAATAGTAAGTACCC |
| *STAT5B* Y665F | TCGTCGGCAGCGTCAGATGTGTATAAGAGACAG | TCAGAATGCGAACATTGTTAC | GTCTCGTGGGCTCGGAGATGTGTATAAGAGACAG | CATGTTTAGAATGTGATTGTTCTG |
| *STAT5B* Q177P | TCGTCGGCAGCGTCAGATGTGTATAAGAGACAG | TCCGAATGGAAAGTGTGC | GTCTCGTGGGCTCGGAGATGTGTATAAGAGACAG | CAGATCAACCAGACGTTTGAG |

**Supplemental Table 15. Hashing antibodies used for sample multiplexing.**

| **Antibody** | **Source** | **Identifiers** |
|---|---|---|
| TotalSeq™-A0251 anti-human Hashtag 1 Antibody; Clones LNH-94; 2M2 | BioLegend | Cat# 394601;<br>RRID: AB_2750015 |
| TotalSeq™-A0252 anti-human Hashtag 2 Antibody; Clones LNH-94; 2M2 | BioLegend | Cat# 394603;<br>RRID: AB_2750016 |
| TotalSeq™-A0253 anti-human Hashtag 3 Antibody; Clones LNH-94; 2M2 | BioLegend | Cat# 394605;<br>RRID: AB_2750017 |
| TotalSeq™-A0253 anti-human Hashtag 4 Antibody; Clones LNH-94; 2M2 | BioLegend | Cat# 394607;<br>RRID: AB_2750018 |
| TotalSeq™-A0253 anti-human Hashtag 5 Antibody; Clones LNH-94; 2M2 | BioLegend | Cat# 394609;<br>RRID: AB_2750019 |

| | | |
|---|---|---|
| TotalSeq™-A0253 anti-human Hashtag 6 Antibody; Clones LNH-94; 2M2 | BioLegend | Cat# 394611;<br>RRID: AB_2750020 |
| TotalSeq™-A0253 anti-human Hashtag 7 Antibody; Clones LNH-94; 2M2 | BioLegend | Cat# 394613;<br>RRID: AB_2750021 |
| TotalSeq™-A0253 anti-human Hashtag 8 Antibody; Clones LNH-94; 2M2 | BioLegend | Cat# 394615;<br>RRID: AB_2750022 |

**Supplemental Figure Legends**

**Supplemental Figure 1. Mutational signature analysis of index patient blood. (A)** Mutational spectrum of mosaic blood-restricted single nucleotide variants showing the relative contribution of each type of base substitution, using the *MutationalPatterns* package in R. **(B)** Contribution of mutational signatures to the patient mutational profile using strict refitting, using the *MutationalPatterns* package in R. **(C)** 96 trinucleotide profile showing relative contribution of mosaic single nucleotide variants, further divided by context.

**Supplemental Figure 2. *STAT5B* variant allele frequencies (VAFs) in disease control PBMCs. (A)** T large cell granular lymphocytic leukemia (T-LGLL) patients with mosaic *STAT5B*$^{Y665F}$ variants. Left and right panels each represent unique patients. **(B)** Growth hormone insensitivity patient with autosomal dominant gain of function *STAT5B*$^{Q177P}$ variant.

**Supplemental Figure 3. Clustering and gene set enrichment analysis (GSEA) details of scRNAseq analysis. (A)** Expression of canonical markers for key immune cell populations (left), supporting cell cluster identity annotations (right). **(B)**. GSEA statistical details of key interferon and STAT related pathways, in patient monocytes (left) and CD8 T cells (right), including false discovery rate (FDR) adjusted q-values and family-wise error rate (FWER) adjusted p-values.

**Supplemental References**


3. Neven Q, Boulanger C, Bruwier A, et al. Clinical Spectrum of Ras-Associated Autoimmune Leukoproliferative Disorder (RALD). *J. Clin. Immunol.* 2021;41(1):51–58.
5. Ma CA, Xi L, Cauff B, et al. Somatic STAT5b gain-of-function mutations in early onset nonclonal eosinophilia, urticaria, dermatitis, and diarrhea. *Blood*. 2017;129(5):650–653.
6. Eisenberg R, Gans MD, Leahy TR, et al. JAK inhibition in early-onset somatic, nonclonal STAT5B gain-of-function disease. *J. Allergy Clin. Immunol. Pract.* 2021;9(2):1008–1010.e2.
7. Ding F, Wu C, Li Y, et al. A case of hypereosinophilic syndrome with STAT5b N642H mutation. *Oxf Med Case Reports*. 2021;2021(1):omaa129.
15. Benjamin D, Sato T, Cibulskis K, et al. Calling Somatic SNVs and Indels with Mutect2. *bioRxiv*. 2019;861054.
23. Rajala HLM, Eldfors S, Kuusanmäki H, et al. Discovery of somatic STAT5b mutations in large granular lymphocytic leukemia. *Blood*. 2013;121(22):4541–4550.
24. Manders F, Brandsma AM, de Kanter J, et al. MutationalPatterns: the one stop shop for the analysis of mutational processes. *BMC Genomics*. 2022;23(1):134.
25. Li H, Durbin R. Fast and accurate short read alignment with Burrows-Wheeler transform. *Bioinformatics*. 2009;25(14):1754–1760.
26. Danecek P, Bonfield JK, Liddle J, et al. Twelve years of SAMtools and BCFtools. *Gigascience*. 2021;10(2.):
27. Knaus BJ, Grünwald NJ. vcfr: a package to manipulate and visualize variant call format data in R. *Mol. Ecol. Resour.* 2017;17(1):44–53.
28. Wickham H, Averick M, Bryan J, et al. Welcome to the tidyverse. *J. Open Source Softw.* 2019;4(43):1686.
29. Masle-Farquhar E, Jackson KJL, Peters TJ, et al. STAT3 gain-of-function mutations connect leukemia with autoimmune disease by pathological NKG2Dhi CD8+ T cell dysregulation and accumulation. *Immunity*. 2022;55(12):2386–2404.e8.
30. R Core Team. R: A language and environment for statistical computing. 2022.
31. Hao Y, Hao S, Andersen-Nissen E, et al. Integrated analysis of multimodal single-cell data. *Cell*. 2021;184(13):3573–3587.e29.
32. McCarthy DJ, Campbell KR, Lun ATL, Wills QF. Scater: pre-processing, quality control, normalization and visualization of single-cell RNA-seq data in R. *Bioinformatics*. 2017;33(8):1179–1186.
33. Butler A, Darby C, Hao Y, Hoffman P, Satija R. Azimuth: A Shiny App Demonstrating a Query-Reference Mapping Algorithm for Single-Cell Data. 2022.
34. Breda J, Zavolan M, van Nimwegen E. Bayesian inference of gene expression states from single-cell RNA-seq data. *Nat. Biotechnol.* 2021;39(8):1008–1016.
35. Ritchie ME, Phipson B, Wu D, et al. limma powers differential expression analyses for RNA-sequencing and microarray studies. *Nucleic Acids Res.* 2015;43(7):e47.
36. Subramanian A, Tamayo P, Mootha VK, et al. Gene set enrichment analysis: a knowledge-based approach for interpreting genome-wide expression profiles. *Proc. Natl. Acad. Sci. U. S. A.* 2005;102(43):15545–15550.
37. Hunter. Matplotlib: A 2D Graphics Environment. 2007;9:90–95.
38. Xie W, Fan G, Wiszniewska J, Press, Richard D., Yang F. Histopathological and molecular findings in a patient with Ras-associated autoimmune leukoproliferative disorder. *Human Pathology Reports*. 2022;29:300670.
39. Papa R, Rusmini M, Schena F, et al. Type I interferon activation in RAS-associated autoimmune leukoproliferative disease (RALD). *Clin. Immunol.* 2021;231:108837.
40. Ziv A, Dardik R, Yacobovich J, et al. Atypical Presentations of Pediatric-Acquired Thrombotic Thrombocytopenic Purpura. *J. Pediatr. Hematol. Oncol.* 2024;
41. Kurita D, Shiba N, Ohya T, et al. Severe RAS-Associated Lymphoproliferative Disease Case with Increasing αβ Double-Negative T Cells with Atypical Features. *J. Clin. Immunol.* 2023;43(8):1992–1996.
42. Tang Y, Wang H, Zhao H, Jin S, Wu J. RAS-associated Autoimmune Leukoproliferative Disease (RALD-KRAS) Consistent with the Clinical Diagnosis of Rosai-Dorfman

Disease: A 15-year Follow-up. *J. Clin. Immunol.* 2024;44(5):123.
43. Blanchard-Rohner G, Ragotte RJ, Junker AK, et al. Idiopathic splenomegaly in childhood and the spectrum of RAS-associated lymphoproliferative disease: a case report. *BMC Pediatr.* 2021;21(1):45.

#### Supplemental Figure 1

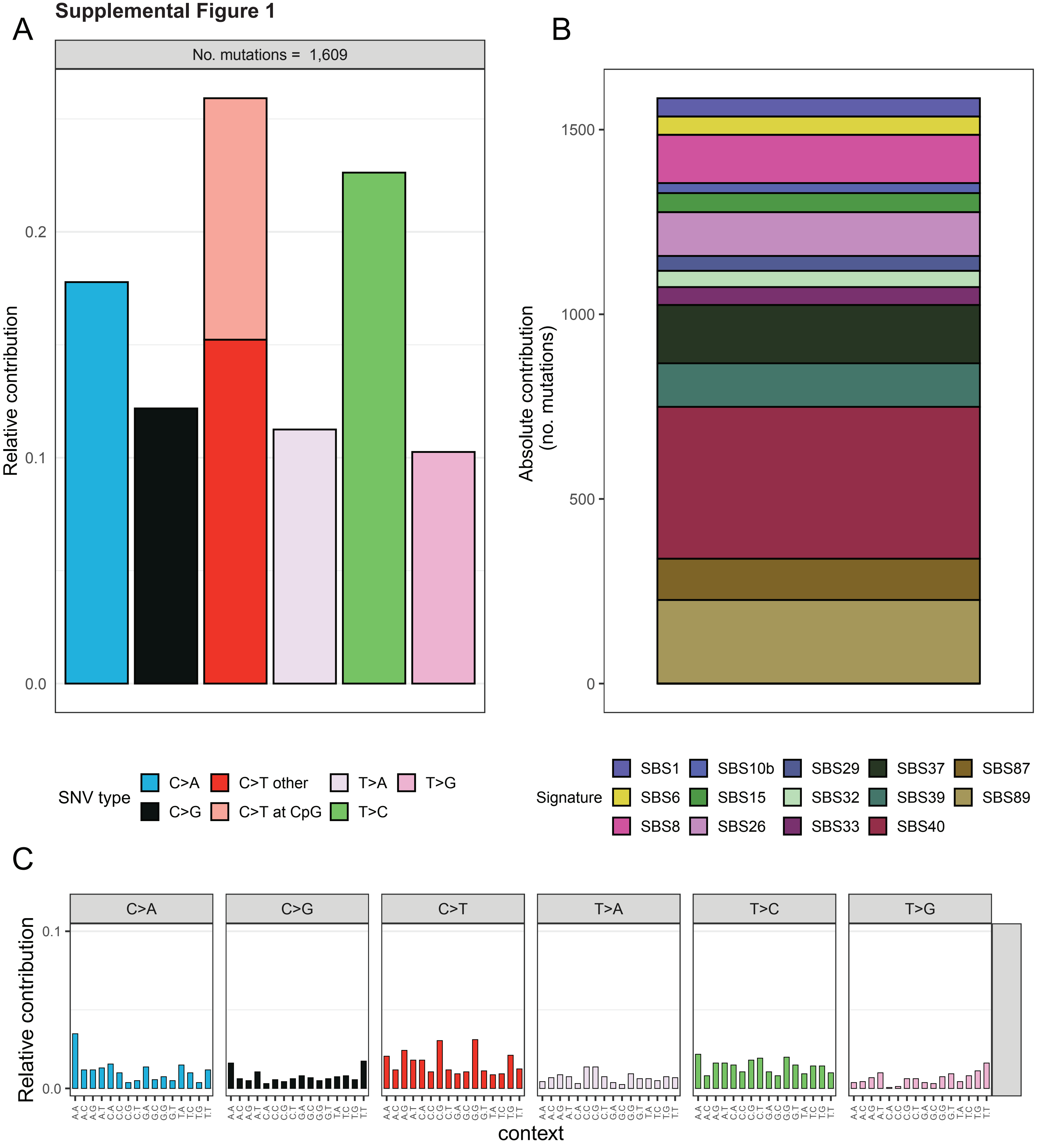

#### Supplemental Figure 2

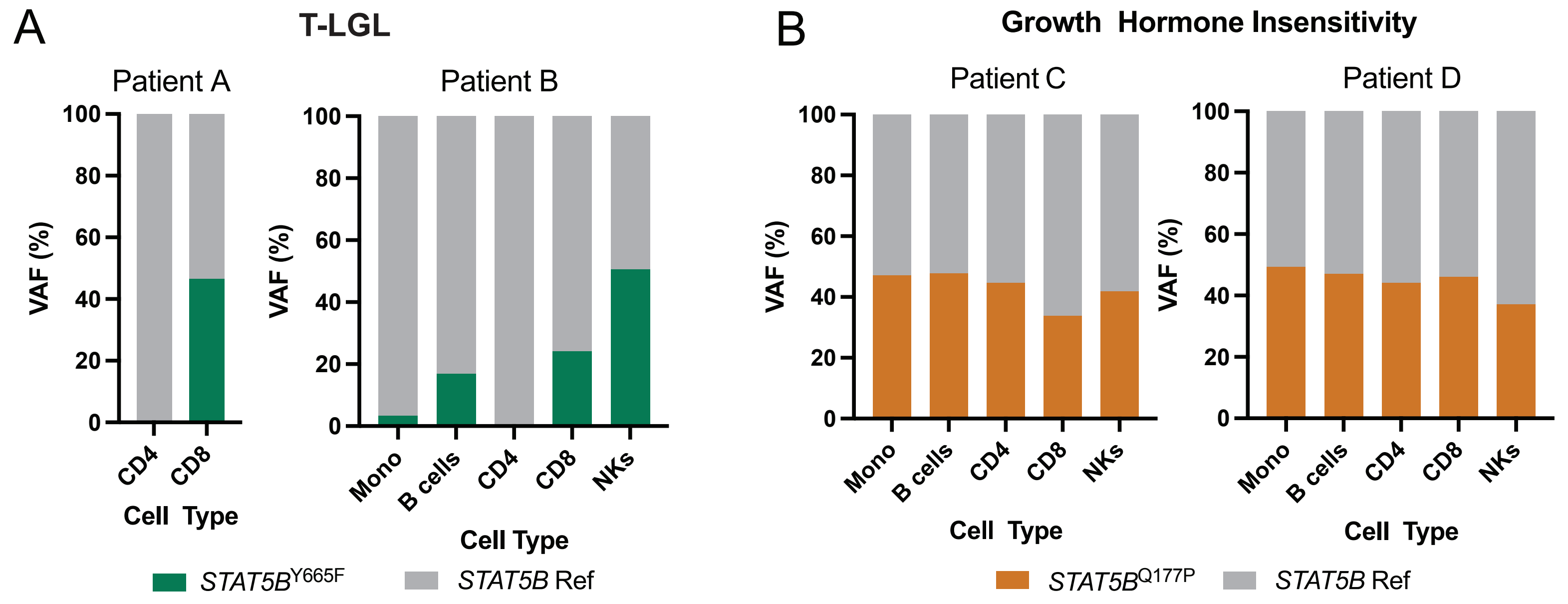

### Supplemental Figure 3

A

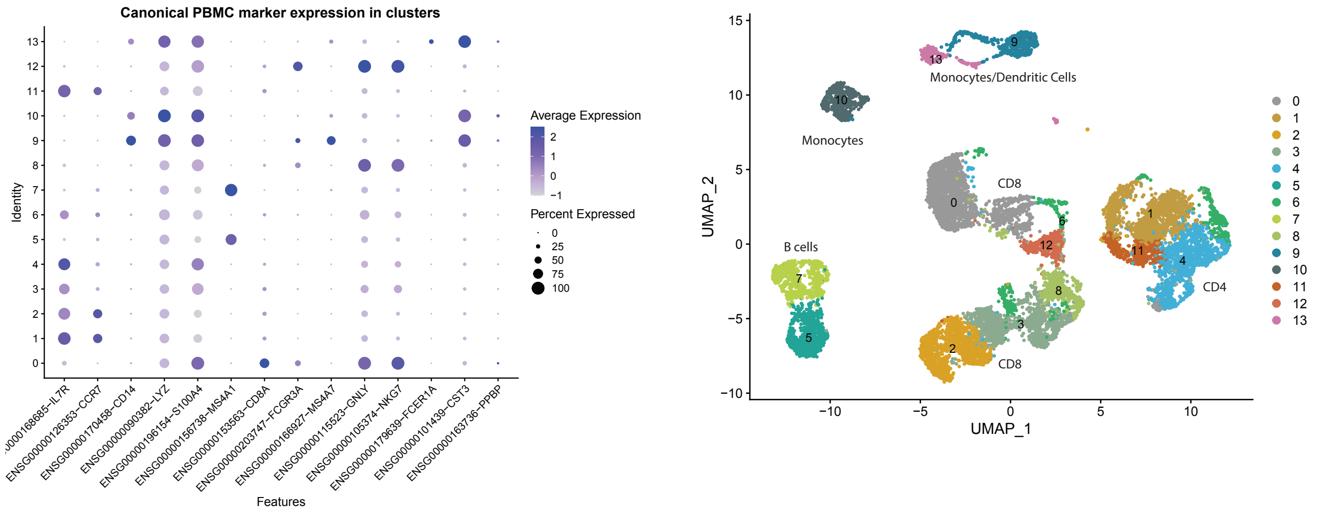


B

#### Monocytes

##### Interferon_Alpha_Response

| | |
|---|---|
| Dataset | IvsH |
| Phenotype | NoPhenotypeAvailable |
| Upregulated in class | na_pos |
| GeneSet | HALLMARK_INTERFERON_ALPHA_RESPONSE |
| Enrichment Score (ES) | 0.55055785 |
| Normalized Enrichment Score (NES) | 1.9172736 |
| Nominal p-value | 2.1486892E-4 |
| FDR q-value | 9.862733E-4 |
| FWER p-Value | 0.0044 |

##### Intereferon_Gamma_Response

| | |
|---|---|
| Dataset | IvsH |
| Phenotype | NoPhenotypeAvailable |
| Upregulated in class | na_pos |
| GeneSet | HALLMARK_INTERFERON_GAMMA_RESPONSE |
| Enrichment Score (ES) | 0.47685277 |
| Normalized Enrichment Score (NES) | 1.8365889 |
| Nominal p-value | 0.0 |
| FDR q-value | 0.0023945058 |
| FWER p-Value | 0.0132 |

##### IL6_JAK_STAT3_Signaling

| | |
|---|---|
| Dataset | IvsH |
| Phenotype | NoPhenotypeAvailable |
| Upregulated in class | na_pos |
| GeneSet | HALLMARK_IL6_JAK_STAT3_SIGNALING |
| Enrichment Score (ES) | 0.4845279 |
| Normalized Enrichment Score (NES) | 1.6436487 |
| Nominal p-value | 0.006728891 |
| FDR q-value | 0.010028717 |
| FWER p-Value | 0.0973 |

##### IL2_STAT5_Signaling

| | |
|---|---|
| Dataset | IvsH |
| Phenotype | NoPhenotypeAvailable |
| Upregulated in class | na_pos |
| GeneSet | HALLMARK_IL2_STAT5_SIGNALING |
| Enrichment Score (ES) | 0.4006185 |
| Normalized Enrichment Score (NES) | 1.5487349 |
| Nominal p-value | 0.0020764119 |
| FDR q-value | 0.018766258 |
| FWER p-Value | 0.2176 |

#### CD8 T Cells

##### Interferon_Alpha_Response

| | |
|---|---|
| Dataset | IvsH |
| Phenotype | NoPhenotypeAvailable |
| Upregulated in class | na_pos |
| GeneSet | HALLMARK_INTERFERON_ALPHA_RESPONSE |
| Enrichment Score (ES) | 0.7459568 |
| Normalized Enrichment Score (NES) | 2.4451923 |
| Nominal p-value | 0.0 |
| FDR q-value | 0.0 |
| FWER p-Value | 0.0 |

##### Intereferon_Gamma_Response

| | |
|---|---|
| Dataset | IvsH |
| Phenotype | NoPhenotypeAvailable |
| Upregulated in class | na_pos |
| GeneSet | HALLMARK_INTERFERON_GAMMA_RESPONSE |
| Enrichment Score (ES) | 0.691588 |
| Normalized Enrichment Score (NES) | 2.4299772 |
| Nominal p-value | 0.0 |
| FDR q-value | 0.0 |
| FWER p-Value | 0.0 |

##### IL6_JAK_STAT3_Signaling

| | |
|---|---|
| Dataset | IvsH |
| Phenotype | NoPhenotypeAvailable |
| Upregulated in class | na_pos |
| GeneSet | HALLMARK_IL6_JAK_STAT3_SIGNALING |
| Enrichment Score (ES) | 0.5998373 |
| Normalized Enrichment Score (NES) | 1.9344684 |
| Nominal p-value | 0.0 |
| FDR q-value | 1.2063306E-4 |
| FWER p-Value | 7.0E-4 |

##### IL2_STAT5_Signaling

| | |
|---|---|
| Dataset | IvsH |
| Phenotype | NoPhenotypeAvailable |
| Upregulated in class | na_pos |
| GeneSet | HALLMARK_IL2_STAT5_SIGNALING |
| Enrichment Score (ES) | 0.45528933 |
| Normalized Enrichment Score (NES) | 1.6050044 |
| Nominal p-value | 2.1184197E-4 |
| FDR q-value | 0.009728848 |
| FWER p-Value | 0.1322 |
